# Opinions of the UK general public in using artificial intelligence and “opt-out” models of consent in medical research

**DOI:** 10.1101/2024.12.13.24318727

**Authors:** William Heseltine-Carp, Mark Thurston, Michael Allen, Daniel Browning, Megan Courtman, Aishwarya Kasabe, Emmanuel Ifeachor, Stephen Mullin

## Abstract

**Background:** Due to its complexity, Artificial Intelligence often requires large, confidential clinical datasets. 20-30% of the general public remain sceptical of Artificial Intelligence in healthcare due to concerns of data security, patient-practitioner communication, and commercialisation of data/models to third parties. A better understanding of public concerns of Artificial Intelligence is therefore needed, especially in the context of stroke research.

**Aims:** We aimed to evaluate the opinion of patients and the public in acquiring large clinical datasets using an “opt-out” consent model, in order to train an AI-based tool to predict the future risk of stroke from routine healthcare data. This was in the context of our project ABSTRACT, a UK Medical Research Council study which aims to use AI to predict future risk of stroke from routine hospital data.

**Methods:** Opinions were gathered from those with lived experience of stroke/TIA, caregivers, and the general public through an online survey, semi-structured focus groups, and 1:1 interviews. Participants were asked about their perceived importance of the project, the acceptability of handling deidentified routine healthcare data without explicit consent, and the acceptability of acquiring these data via an opt-out model of consent model by members within and outside of the routine clinical care team.

**Results:** Of the 83 that participated, 34% of which had a history of stroke/TIA. Nearly all (99%) supported the project’s aims in using AI to predict stroke risk, acquiring data via an opt-out consent model, and the handling of pseudonymized data by members within and outside of the routine clinical care team.

**Conclusion:** Both the general public and those with lived experience of stroke/TIA are generally supportive of using large, de-identified medical datasets to train AI models for stroke risk prediction under an opt-out consent model, provided the research is transparent, ethically sound, and beneficial to public health.

## Introduction

Patient and public involvement (PPI) describes the process of actively engaging patients, carers and the general public in designing, conducting and disseminating research. There are a number of benefits of PPI. Firstly, it provides an opportunity to tailor research to the needs and values of patients. Secondly, it ensures research is conducted in a way agreed to be ethical and acceptable. Thirdly, it enhances trust from patients and the public in research by empowering them to take ownership and an active role in research. Finally, it enables better dissemination and accessibility of results.

Increasingly, evidence of PPI input is often required by funders and ethical review boards. This further highlights the importance in researchers undertaking such work in order to produce ethical and high-quality research.^1^

As healthcare records and investigations become increasingly digitalised, opportunities emerge to analyse these data to improve clinical care delivery and answer research questions. AI techniques such as machine learning (ML) have the capacity to analyse high-dimensional data with thousands of features, so are often better-suited to the analysis of large, complex, multi-faceted datasets than traditional statistical techniques.

It is likely that AI will continue to accelerate medical research and transform clinical practice. However, with the significant capabilities that AI offers, come the need for appropriate stewardship and responsibility. Approximately 20-30% of individuals remain sceptical of the use of AI models in medical research.^2^ Due to its complexity, large datasets are often required to train AI models. Since it would be impractical to gain consent from each individual, data is often de-identified, and individuals excluded on an opt-out basis. Hence, much of this scepticism relates to data acquisition, storage and security of data. Naturally, societal impacts of AI have also been raised, including job security, effective communication with patients and sale of data/models to external organisations.^3^

The use of routine healthcare data in medical research presents new challenges as well as major opportunities. In order to assure that the data used is representative, it must be as comprehensively collated as possible. Additionally, in order to undertake such research in an ethical and compliant manner, it is essential to demonstrate a valid medical purpose and confirm support for use of data without consent amongst key stakeholders, in particular, those with lived experience of the disease and (where the use of ‘control’ data is required) the public at large. High quality PPI work is therefore increasingly an important prerequisite to this type of research.

### Aims

Our group is currently using AI to predict future risk of stroke based on routine hospital data from 120,000 patients (ABSTRACT)^4^. In this article we report the findings of our PPI work for this study, in which we aimed to evaluate the opinion of patients and the public to the following three questions:

1. *Is it acceptable to train AI to predict future risk of stroke from routine hospital data?*
2. *Is it acceptable to acquire and handle large patient datasets using an “opt-out” model of consent?*
3. *Is it acceptable for members both within and outside of the routine clinical care team to have access to these datasets?*

To understand the context of our data handling procedure our protocol is published elsewhere^5^. Figure 1 illustrates the data handling procedure for this study. In brief, cases are identified by the data controller, who is a member of the routine clinical care team (i.e. an individual who would normally have access to this data for clinical purposes). Cases are identified from the Sentinel Stroke National Audit Programme (SSNAP) database^6^, a mandated national audit of stroke cases in the UK, which contains data related to every inpatient UK stroke admission. Based on these data a pseudonymised list of National-Health-Service (NHS) numbers (the unique identifier assigned to every UK citizen) of stroke cases is generated. This pseudonymised list is then shared with data providers, such as NHS hospitals, integrated care boards and GP practices, who then provide data on the cases and generate matched controls for the data controller. The data controller then links these data to form a single, pseudonymised dataset of cases and controls. Participants are then compared against the national opt-out database (NDOO)^7^ and removed accordingly. The dataset is then de-identified by the data controller before being shared with the research team for analysis. At all stages the minimum number of data handlers are given access to the dataset, and data is stored on encrypted NHS computer systems. All data will be handled in compliance with GDPR and University of Plymouth’s “research ethics & integrity policy”^8^ and “research data management policy”^9^.

**figure 1.**
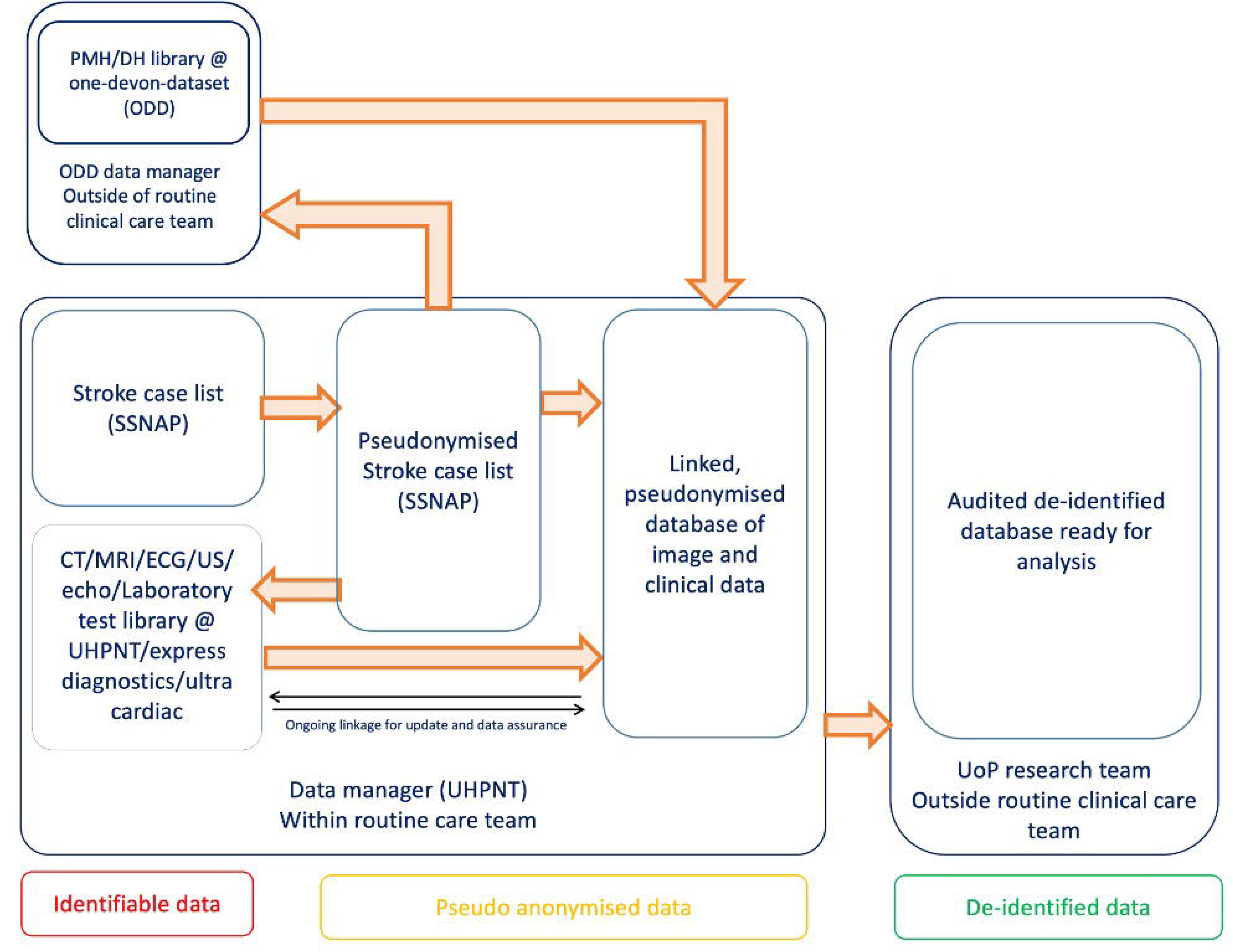

## Methods

To ensure a representative sample, we used a variety of sampling methods. These were an in person/online (hybrid) semi-structured focus group, an online questionnaire and one to one interviewing of inpatients at Derriford Hospital, UK.

### Focus group

Those with lived experiences of stroke/TIA and members of the general public were invited to join two focus groups discussing their opinions on training AI to predict future risk of stroke using routine hospital data. Participants were invited to attend online or in person. This was advertised via social media pages of the University Hospitals Plymouth NHS Trust trust, Stroke Association^10^ and National Institute for Health and Social Care Research (NIHR) South West Peninsula Clinical Research Network (CRN)^11^.

Participants were offered a £20 Amazon voucher^12^ for their time and reimbursed for travel expenses. Registration took place via online survey. Participants were then contacted via email to communicate session details. Prior to the session participants were provided with two documents. The first provided background information on stroke, AI and the roles of PPI (appendix 1). The second provided details on how our study plans to acquire, handle, process and store data (appendix 2).

Each focus group consisted of one 90-minute session and used a semi-structured discussion guide structure. Ten minutes were dedicated to a presentation briefly defining and explaining stroke and use of artificial intelligence in research. A further 10 minutes were dedicated to explaining how our study planned to collect, store and analyse data, in addition to explaining how to opt out of the study. Participants were then asked to discuss three questions in groups of three to four. These were: *“1) Is it acceptable to collect, store and analyse data in this manner for medical research inside and outside the routine clinical care team?”; “2) Has sufficient opportunity for participants to ‘opt out’ of research has been given?”; “3) Is there anything further you would like to add?”*. After each question a consensus was reached and summarised as *“(YES/NO/UNABLE TO REACH CONSENSUS)”*.

### One-to-one interview

Stroke inpatients and family members were invited for a 10-15 minute 1:1 interview on assessing their opinion of using AI to predict future risk of stroke using routine hospital data. For each interview the rationale and methodology of the study was described, followed by details of how the study plans to collect, store and analyse data, and the opportunity to opt out. Participants were then asked the same three questions as above, summarised with a *“YES/NO”* answer.

### Online survey

An online survey was designed using “Qualtrics XM^13^”. Data were obtained from the dates of 9.2.24 to 28.7.24 from 61 participants. Advertisement was done via the study website and social media posts from the local NHS trust. The survey included information on the rationale and methodology of the study, in addition to how data will be collected, stored and analysed. Participants were asked four YES/NO questions and had the option to add further comments to each answer. These consisted of: *“Do you believe these study aims and objectives are important?”; “Do you believe it is appropriate to use NHS data in this way?”; “Do you think this an appropriate way to handle and protect patient data?”; “Under these circumstances, is it appropriate for trained NHS staff who would not normally have access to it (i*.*e not part of the routine care team) to handle data in this way?”; “Is there anything else you would like to see included in the design of this study?”*. Data on prior history of transient-ischaemic-attack(TIA)/ stroke were also collected.

### Storage of data

Focus group and interview sessions were audio recorded and stored as an MP3 file on an encrypted and password protected device. Online survey data were stored on the Qualtrics XM^13^ platform under an encrypted password protected server. Data collection, handling and storage was compliant with guidance outlined by local NHS trust, University of Plymouth^9^ and the General Data Protection Regulation (GDPR)^14^.

### Ethical approval and consent

Written and/or oral consent was obtained from all participants. Ethics and data handling procedures complied with the University of Plymouth’s “research ethics & integrity policy”^8^ and “research data management policy”^9^.

## Results

### Focus group

A total of 16 participants attended the two focus groups. 7 attended online and 9 in person. Mean age was 67. 65% were male. 36% had a history of stroke/TIA and 57% had close friends or family affected by stroke/TIA.7% had no experience of stroke. There were 8 participants for the first focus group and 9 for the second. Two withdrew participation on the day as they did not feel confident in contributing due to cognitive and communication deficits.

1. *Is it acceptable to collect, store and analyse data in this manner for medical research inside and outside the routine clinical care team?*
  A. Within the routine clinical care team? All participants responded with “YES”. Participants felt that the research was worthwhile and that the measures taken to maintain participant confidentiality and protecting personal data were sufficient.
  B. *Outside of the routine clinical care team?* All participants responded with “YES”. However two participants stressed that they would object to the data being used for commercial purposes, particularly in the context of medical insurance, tech and pharmaceutical companies. Instead, they felt the data should be used for research purposes only. Participants did not object to data being shared with researchers from other regions within the UK.
2. *Has sufficient opportunity for participants to ‘opt out’ of research has been given?* All participants felt that sufficient opportunity to opt out was provided via NDOO^7^ and the study website^4^. In order to improve community outreach, three participants suggested opt-out could be advertised using posters in GP practices, supermarkets and post offices. Two participants also suggested creating an additional “opt-in” register, allowing those who had signed up to the NDOO to opt their data into the project.
3. *Is there anything further you would like to add?* Participants wished the project well and many expressed gratitude for the research given their personal experience with stroke. Two participants volunteered to join an oversight committee for further PPI involvement in the project.

Potential impacts for the research were also briefly discussed. This included discussion around designing a clinical trial in which individuals who are identified as high risk of stroke could be treated with placebo vs pharmaceutical and/or lifestyle intervention. All participants felt this would be appropriate and valuable research.

### One-to-one interview

Sixteen individuals were approached in the inpatient setting. Fourteen agreed to participate in a 1:1 interview. Twelve had a history of stroke and 2 cared for someone with stroke. 57% were male. Mean age was 71. Mean interview time was 9 minutes 32 seconds.

1) *Is it acceptable to collect, store and analyse data in this manner for medical research inside and outside the routine clinical care team?*

All participants responded with “YES”. Participants generally felt that handling data in this way was justified by the stated medical purpose, namely to improve stroke care. No participant objected to sharing this type of data outside of the routine clinical care team, however one participant stated a preference for the data to remain within the UK. One participant specified that they would only want data relating to their health to be collected, and not other forms of personal data (e.g. financial).

2) *Has sufficient opportunity for participants to ‘opt out’ of research has been given?*

All participants felt that sufficient opportunity to opt out was provided via NDOO^7^ and the study website^4^. They were satisfied with current methods of advertising the study via local NHS trust social media. Three participants suggested advertising the project via the “stroke association” and “Age UK”.

3) *Is there anything further you would like to add?*

Participants generally used this opportunity to express their support for the study. One participant stressed that they felt the data should be used for academic research only, and not sold/shared with insurance, technology or drug companies.

### Online survey

There were 61 respondents to the online survey. 17% had a history of stroke/TIA and 38% had friends of family affected by stroke (figure 2). 36% had no lived experience of stroke.

**figure 2.**
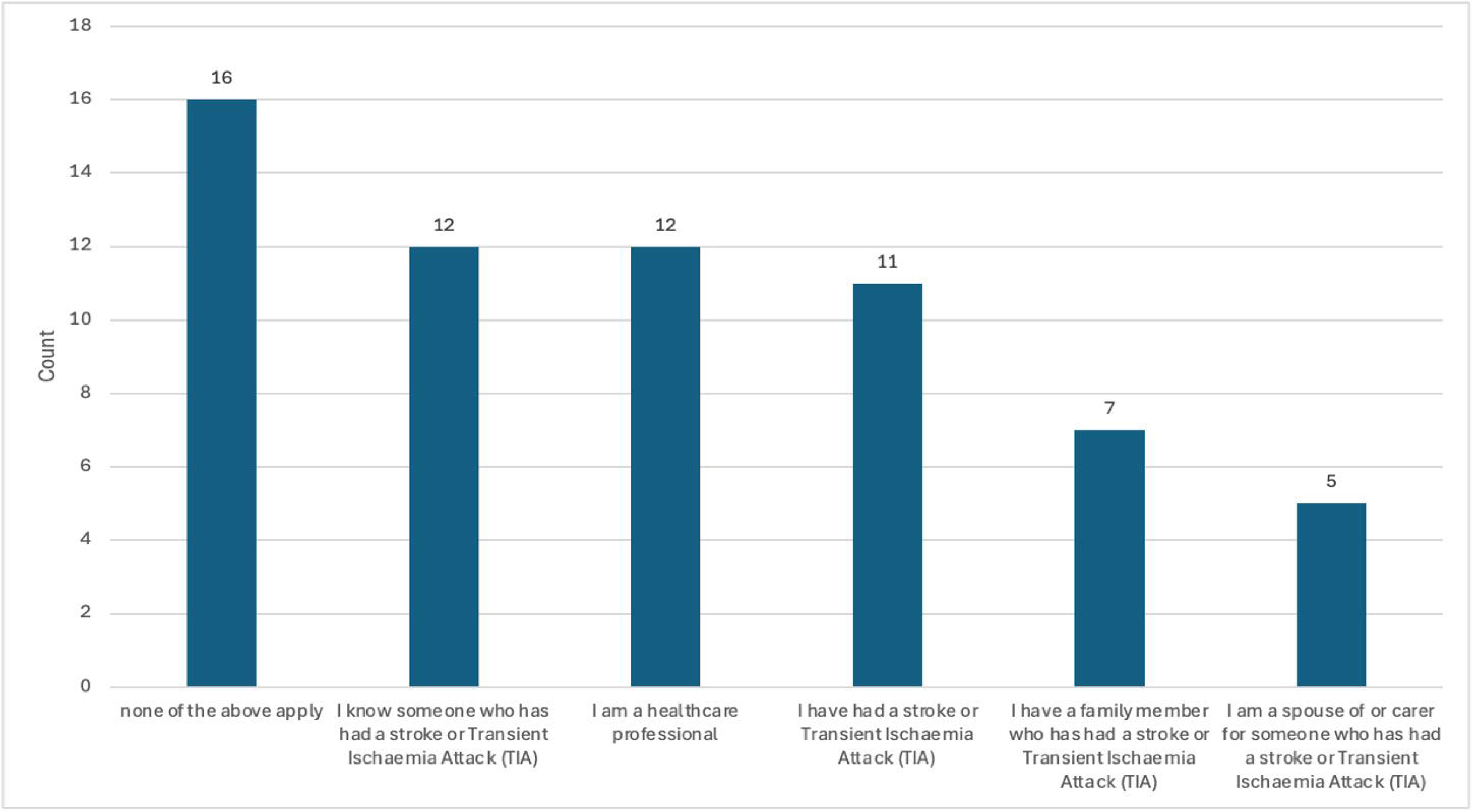

Figure 3 and table 1 illustrate the survey results. Sixty of 61 respondents felt the study aims and objectives were important. Sixty of 61 respondents felt this was an appropriate way to use NHS data. Three participants commented that they were reassured that the data would remain within the NHS or University. Sixty of 61 felt it was appropriate for data to be handled by members within the routine clinical care team. Fifty-nine of 62 felt it was appropriate for data to be handled by members outside of the routine clinical care team. Two participants commented that they felt that the stated medical purpose justified such unsolicited data access. One participant commented that “as long as the fewest number of people have access”. One participant responded “no” to all answers, but did not specify why.

**figure 3.**
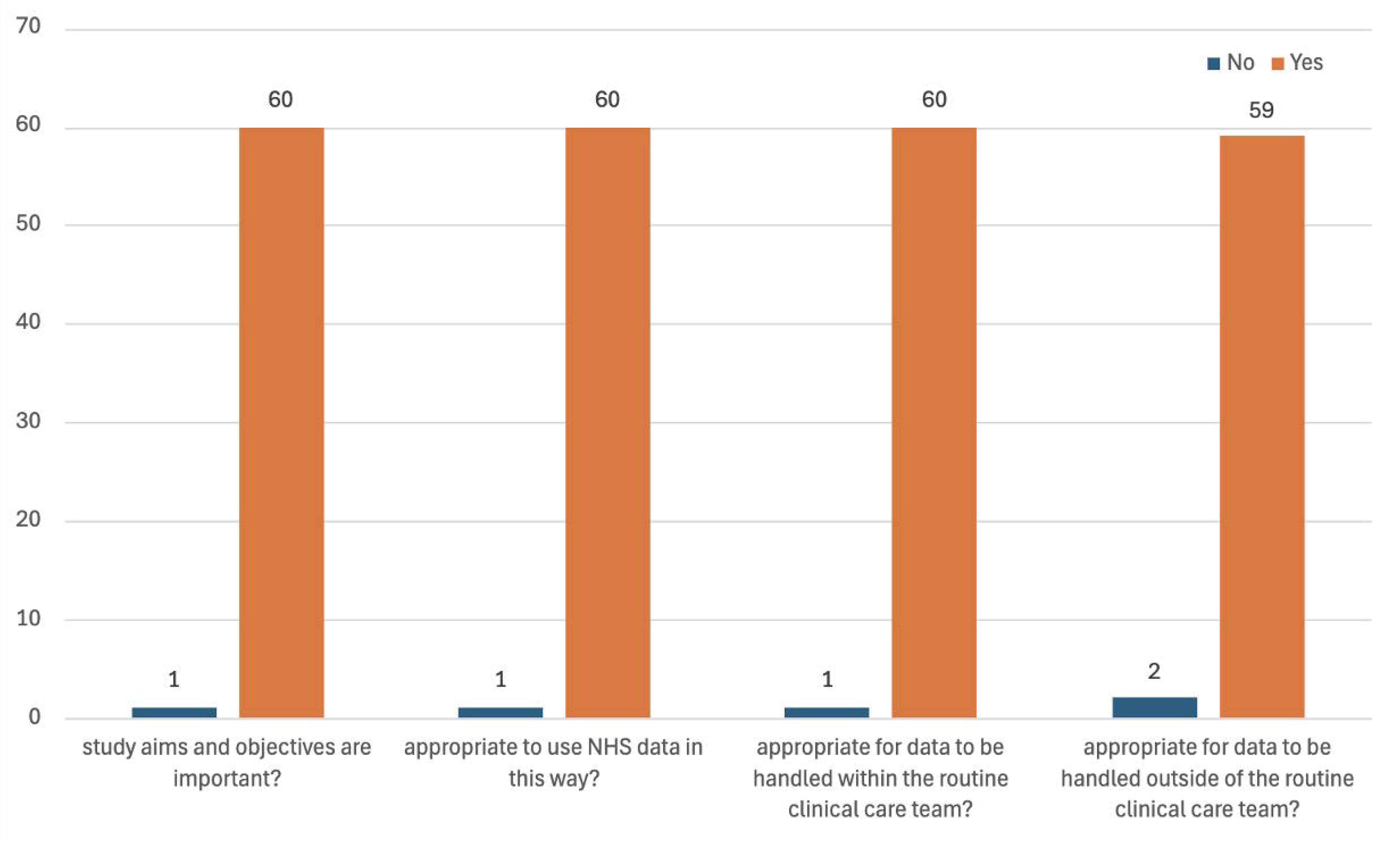

### Amendments to study design based on PPI feedback

Following feedback obtained via our PPI work, three major opportunities for change were identified:

Firstly, our strategy in publicising opting out of our project has mainly focused in the inpatient hospital setting and online social media. However, participants’ feedback identified that those “less computer literate” or healthier populations who have only attended hospital on one-off occasions may not be reached via these platforms. Hence, we have further publicised the project via posters in community spaces such as supermarkets, GP surgeries and post offices to widen our catchment. Based on participant feedback, we have also reached out to “AgeUK” and the “Stroke Association” to advertise the project online and via the post.

Secondly, participants highlighted the importance of transparency in medical research. We have therefore recruited a “study oversight group” to oversee the design, development and potential impacts of the study.

Thirdly, participants felt that many individuals may be unsure of their data sharing preferences and hence suggested creating a local “opt-in” register. A register was therefore created to enable patients and the public to opt their data into the project. The details on how to opt in were advertised via poster, social media and the study website.

## Discussion

In this article we publish the findings of the PPI work for the ABSTRACT study^4^. We evaluated the opinions of a total of 83 volunteers in using AI to predict future risk of stroke from routine healthcare data and in acquiring this data via an opt-out consent model. This was done using a combination of patient interview, focus group and online survey. To our knowledge, no other study has sought to investigate the opinions of the general public and patients in using AI to predict future risk of stroke.

Nearly all participants felt that this was an appropriate use of NHS data and that it was acceptable for the data to be handled by members both in and outside of the routine clinical care team. Nearly all participants also felt that handling these data without gaining explicit patient consent was justified by the stated medical purpose of preventing future stroke. Likewise they supported the use of an opt out system.

Based on our study protocol, participants generally also felt that sufficient opportunity was given to opt out, however several participants suggested advertising study posters in public places such as GP surgeries, supermarkets and post offices. Only one participant (online survey) felt that the study aims were not important and did not feel that this was an appropriate use of NHS data, but did not explain why.

These consensuses were based on several conditions. Firstly, the data would be used for research purposes only, and not sold to third party organisations. Secondly, the data would be handled in compliance with GDPR guidance^14^ and only stored on encrypted NHS and/or university devices. And thirdly, the data would be collected from the minimum number of participants and handled by the minimum number of researchers.

With respect to AI in medical research, these findings are in agreement with several other studies that have demonstrated a favourable public opinion in using AI in medical research, but were sceptical about the accuracy, accountability and ethico-legal implications of AI in healthcare^3,15–17^. The common concerns that were raised in both this and other studies included cybersecurity, data leaks and the sale of data to commercial bodies (which might impact on health insurance, mortgage, etc.)^3,16–18^.

However, our cohort generally felt that the potential benefits of this research outweighed these risks, assuming data handling was compliant with GDPR and the data would not be sold to commercial bodies. This is consistent with a recent government survey that identified public concerns around AI could be mitigated with public education, project transparency and adherence to ethical, legal and governance frameworks^3,18^.

Previous research has also highlighted concerns of AI dehumanising medical care and threatening job security^2,15^. Since the accuracy of an AI algorithm is largely dependent on the quality of the training data, concerns have also been raised about the reliability of AI driven medical decision making, especially in minority groups who may be underrepresented in the training data set^19^. None of our participants volunteered these concerns, however they were not explicitly discussed.

With regards to consent, a large body of research has evaluated the public opinion of adopting an opt-out strategy in consenting for medical research^20–22^. Our findings are consistent with this research in that both the general public and patients are generally supportive of opt-out models of consent. However, again, this is often on condition that projects maintain transparency, adhere to ethical and legal frameworks (such as GDPR) and that it is perceived that the research will provide clear health benefits^23–25^. The latter of which is highly influenced by public and patient education, in addition to their trust in the institution performing the research^26^.

### Limitations

Our project was subject to several limitations. Firstly, when recruiting for a focus group, many stroke patients were unable to attend in person or virtually. Some also did not feel confident in contributing due cognitive or communication deficits. Consequently, those with advanced disability or without access to a computer may be underrepresented.

However, we attempted to mitigate this underrepresentation by performing 1:1 interviews with stroke patients in the hospital setting.

Secondly, our cohort encompasses a total of only 83 participants from the Cornwall and South Devon, UK area, and hence these findings may not be generalisable to the entire UK/world population.

Thirdly, given that the participants generally held favourable views of our methods, little information was obtained about why individuals may object to using AI in medical research or adopting an opt-out consent model, making it difficult to understand ways to mitigate these barriers.

Finally, we assessed these views in the context of our ongoing project, ABSTRACT, which aims to use AI to predict future risk of stroke from routine hospital data^4^. Hence, the views highlighted here may not be representative of views held for similar or other AI based medical research projects. Assessment of project risk vs benefit and transparency is subjective, and may vary between communities, highlighting the need for project specific PPI work.

### Conclusion

We believe that this research identifies two important findings. Firstly, both the general public and those with lived experience of stroke appear supportive of the use of large, de-identified medical datasets to train AI models to predict future risk of stroke.

Participants also supported the acquisition of these datasets via an “opt-out” consent model, but this was under the condition that the research process is transparent, adherent to ethical and legal frameworks, aims to provide clear health benefits and sufficient opportunity is given to opt out.

### Future research

We believe these findings offer an important perspective for ethical reviewers when considering medical research involving AI and opt-out models of consent. Although public support appears favourable, an enhanced understanding of why people may object to AI in medical research is needed, in order to further AI governance and improve public trust. We also accept that the sample of this research was not representative of the diversity of the UK, therefore additional work is required in communities where it is known there is a higher level of distrust of medical research involving large datasets^27^. This work is planned for subsequent phases of the ABSTRACT project.

## Supporting information

Table 1: Survey responses (comments)

## Data Availability

All data produced in the present study are available upon reasonable request to the authors

