## Supplementary material for "Opinions of the UK general public in using artificial intelligence and “opt-out” models of consent in medical research": Table 1: Survey responses (comments)

| **Study aims and objectives are important?** | **Appropriate to use NHS data in this way?** | **Appropriate for data to be handled within the routine clinical care team?** | **Appropriate for data to be handled outside of the routine clinical care team?** |
| --- | --- | --- | --- |
| I think it is very important to find AI tools to identify more and more conditions. AI can be very accurate and from patient data new studies can be done. | I think it is a good way to analyse and use patient data. | I think it is a very good way to manage patients opt out and consent. Historical data don't need patient consent anyway and the data will be de-identified. | Without this progress would be hindered |
| It's always better to be prepared as much as possible so identifying previously unknown risk factors for stroke is a very good plan! | Anything that can help detect risks before any TIA or stroke is both good for patient/family and NHS. | No it seems covered . Information Governance is important but you need the data to avoid future health issues . | The purpose justifies this access |
| I think the objectives are a great idea minimising risk of stroke is important | Sounds like it will stay within the NHS, in which case this is fine. |  | as long as the fewest number of people have access |
| Anything that can prevent people from having a stroke would be an amazing thing. | if we can reduce stroke it will prevent untold misery |  | I would be concerned about patient confidentiality |
| seems like an important project | yes. I am reassured that it stays within the NHS |  |  |
| very important | I am sure the data will be suitably protected and anonymised |  |  |
| I support all medical studies that seek to prevent health conditions and aim to increase the health of the population and save resources used for recovery | Think it's a great proactive idea. Mum had a major stroke at 43 which could have been prevented . I'm keen to prevent myself having one. |  |  |
|  | Seems reasonable given the study aims |  |  |
| **Table 1: Survey responses (comments)** | | | |
