## Appendix 1 for "Opinions of the UK general public in using artificial intelligence and “opt-out” models of consent in medical research"

Chief investigator: Dr Stephen Mullin

Room N6
ITTC Building Plymouth Science Park
Derriford
Plymouth
PL6 8BX

### PARTICIPANT INFORMATION SHEET

#### Patient and Public involvement: The use of routine hospital data in clinical research.

We'd like to invite you to take part in our focus group. This is to discuss the ethics of using anonymised routine hospital data in research, without gaining elicit patient consent. Before you decide, it is important that you understand why the research is being done and what it would involve for you. Please take time to read this information and discuss it with others if you wish. If there is anything that is not clear, or if you would like more information, please ask us.

#### Background

*What is a stroke?*

Stroke can be a highly debilitating illness that affects approximately 100,000 individuals in the UK per year. Stroke occurs when blood flow to the brain is distributed resulting in death of brain cells in the affected area. Around 85% of strokes are caused by a blood clot in a major vessel supplying the brain (ischaemic stroke) and around 15% are caused by a bleed on the brain (haemorrhagic stroke).

This can cause symptoms such as weakness and/or loss of sensation down one side of the body, problems with memory, language, self awareness and vision. In most cases people are left with some form of long lasting deficit, with only 25% returning to their pre-stroke level of functioning. Consequently stroke survivors often require major changes to their lifestyle with many requiring additional care and support.

*What are the risk factors for stroke?*

We know that many risk factors can contribute towards stroke, such as high blood pressure, high cholesterol, diabetes and an irregular heart rhythm (atrial fibrillation).

We therefore prescribe medications to help lower blood pressure, cholesterol, and blood thinning medications to help reduce the risk of someone having a stroke.

Despite treatment many people go on to have a stroke. From previous research we also know that around 1 in 3 people who are admitted to hospital with stroke have no clear risk factors or cause for the stroke, meaning their stroke is unexplained.

This might mean that there many risk factors for stroke that we don't yet know about, and that further research is required to identify these novel risk factors.

#### Why are we conducting this research?

*Rationale*

As mentioned, much of our current management of stroke is based on prescribing medications that target peoples risk factors for stroke and therefore reducing the risk of them having a stroke in the future. However, many people without these risk factors still go on to develop stroke.

There is therefore a need to better understand what puts someone at risk of having a stroke and how we can accurately predict the probability of someone having a stroke.

*Aims*

Our project therefore has two aims:

1. To build a computer programme capable of accurately predicting someone's risk of having a stroke in the future.
2. To use this programme to identify new, previously unknown risk factors for stroke.

*Intended benefits for patients*

We hope this research will eventually lead to research that provides a number of benefits, including:

1. Identifying more people deemed high risk of having a stroke. This could eventually be used to start more people on preventative treatment to reduce their risk of stroke in the future.
2. Identify new risk factors for stroke, which might help in the discovery of new treatments for stroke.

#### How do we plan on achieving this?

To do this we plan on using a powerful computer tool called artificial intelligence (AI). AI is a form of analysis in which machines process, perceive and infer information. In recent years it has successfully led to new discoveries across a wide range of industries.

The caveat is that many AI projects require extremely large datasets. In theory the larger the datasets, the more accurate stroke prediction programme we will be able to build. It may also increase our chances in finding new risk factors for stroke.

*How do we plan on building this large dataset?*

When someone visits a hospital, they often undergo a series of routine medical tests, including physical examination, blood tests, heart scans and scans of their brain or other parts of the body.

Every day, hundreds of people visit Derriford hospital and thousands of investigations are performed. This means that a huge amount of data is held on hospital servers.

Within University Hospitals Plymouth NHS Trust (UHPNT) and the University of Plymouth (UoP), there is an increasing focus on undertaking research using this routinely collected data collected as part of 'routine clinical care' i.e. the investigations that are routinely performed when someone visits hospital.

In our project we are looking at the data of 10,000 stroke patients and 80,000 patients who haven't had a stroke, from Derriford hospital over the last 10 years. We will be looking at information, such as their brain scans, heart scans, past medical history, and physical observations.

We will then use this information to build a computer programme that will be able to predict someone's risk of having a stroke.

*But what about gaining consent from people to use their data?*

In most research projects participants are talked through the study and sign a written consent form to allow researchers to use their data.

We'll be performing analysis of data from approximately 90,000 patient records. Gaining consent from such a large number of patients is impractical. Gaining consent from only a portion who are able to be contacted to give consent is likely to distort results, leading potentially to incorrect conclusions being drawn. This means that it is necessary to make use of anonymised data without consent of the patients.

We understand that not everyone will be happy about having their data used in this way. That's why we want to put together a focus group to discuss some of the issues that people might have in using data in this way and ways we can overcome this.

#### Why have I been invited?

In order to do so in an ethically/legally appropriate manner and to maintain public trust, it is necessary to get feedback on the design of such work from members of the general public and those affected by a number of medical conditions. This is why we have invited you to our focus group discussion.

You've been invited because we are looking for 10 members of the public and 10 people who have had a stroke in the past to join our focus group. In this focus group we'll discuss the 3 questions which are highlighted below. These questions are designed to explore some of the issues that we might encounter by accessing, analysing and storing people's data without their elicit consent.

The purpose of this session is to get your feedback on how we collect and use this data in a number of ongoing/planned research projects. Your feedback will be incorporated into the design of these projects to ensure we do this in an ethical manner.

#### Do I have to take part?

You don't have to take part. Participation is entirely voluntary, and you will be free to withdraw from the focus group at any time.

#### What will happen to me if I decide to take part?

We would like to invite you to a focus group session to discuss the below questions relating to our research. The session will last approximately 1hr 30 mins on 28/02/24.

This would ideally be in person, but there will also be the option to join over video call should you choose.

The day will consist of approximately 9 other stroke survivors (total of 10) and begin with a 10 minute presentation summarising our project and the way the data is handled. This will be followed by 10 minutes of questions on the presentation or any points from the pre-reading that need clarification.

We will then review each of the 3 questions below and break out into small groups of 3 or 4, where you will discuss each question amongst your group. You will be asked to decide on a consensus for each question.

At the end of each question we will conclude a summary answer (YES/NO/UNABLE TO REACH CONSENSUS), together with a short summary of the discussion. A transcript of the call will be produced.

You are more than welcome to bring a relative, friend or carer with you to engage with the discussion.

#### Session details

Details of the session are outlined as followed:

*Prior to arrival*

We ask you to review the 'pre-read' documents below prior to the start of the session. This consists of five questions addressing potential issues with our research. Below each question is our current plan to address each issue. These are difficult concepts so don't worry if you don't fully understand each point. There will be a chance to ask questions and clarify points on the day.

*On the day*

10:00 - 10:10: Arrival and welcome tea and biscuits.

10:10 - 10:20: Presentation on our project and the ways in which we plan on handling data.

10:20 - 10:30: Questions on the presentation and pre-reading.

10:30 - 10:45: Break

10:45 - 11:30: Break out group discussions for each question.

11:30: Concluding remarks

#### Questions for the day

We will discuss the following questions around handling data on the day.

Prior to the day you will be given a pre-reading information pack. This includes information relevant to each question and how we plan to address the issue that the question is highlighting.

**1) Is it acceptable to collect, store and analyse data in this manner for medical research within?**

**2) Has sufficient opportunity for participants to ‘opt out’ of research has been given?**

**3) Is there anything further you would like to add?**

#### Venue and location:

The focus group will be held on 28/02/24 From 10:00 to 11:30 at the following address:

ITTC Building Plymouth Science Park
Derriford
Plymouth
PL6 8BX

#### Will I be reimbursed for taking part?

We will be able to cover your travel costs up to £25. Please provide evidence of your travel receipt if using public transport. If you are travelling by car we are able to reimburse you up to 45 pence per mile.

We would also like to offer you a £20 amazon voucher at the end of the day to thank you for your time.

#### Are there any possible disadvantages or risks from taking part?

We don’t anticipate that involvement in the focus group would pose any risk to yourself or others. However, in some cases we may talk about your experiences with stroke and how it has affected your life. We recognise that for some people this might be distressing or difficult to talk about.

#### Will my taking part in the study be kept confidential?

*Your consent and confidentiality*

At the start of the session, we will ask you to sign a consent form to allow us to record the discussions throughout the day, create a report summarising the discussions and publish these findings.

These recordings will be stored on an encrypted device and be made only available to members of the study team. Once a report has been created and published the recordings will be destroyed.

All participants will be anonymised, and no participant identifiable information will be stored or published.

To ensure confidentiality we ask you to avoid identifying yourself or other members of the group.

#### What will happen to my data?

*Research is a task that we perform in the public interest.* University hospital Plymouth NHS trust*, as sponsor, is the data controller. This means that we, as* University hospital Plymouth NHS *researchers, are responsible for looking after your information and using it properly. We will use the minimum personally-identifiable information possible. We will keep identifiable information about you for 1 year after the focus group has finished. We will store the anonymised research data and any research documents with personal information, such as consent forms, securely at the* University hospital Plymouth NHS trust *for 1 year after the end of the study.*

#### What will happen if I don't want to carry on with the study?

Participation is voluntary and participants may change their minds at a later stage.

Withdrawal will not affect the care you receive from any relevant service (e.g. for patients, from the NHS).

If you withdraw from the study, we will destroy all your identifiable information, but will use the data collected up to your withdrawal, unless you specify otherwise.

#### What will happen to the results of this study?

The results of the focus group will be used to inform how we collect, store and analyse data without gaining elicit patient consent in the future.

We will summarise the findings and discussions of the focus group in a written report and anticipate publishing this in an academic journal and presenting our findings at academic conferences.

No participant identifiable information will be included in the written report, publication, or presentation.

#### Complaints

if you wish to complain, or have any concerns about any aspect of the way you have been approached or treated during the course of this study, you should contact Dr William Heseltine-Carp at or Dr Stephen Mullin at

You will also be able to contact the Patient Advice and Liaison Service (PALS) in the first instance (01752 439884).

#### Who is organising and funding the study?

This is a joint project between the University of Plymouth and University hospitals Plymouth NHS trust.

The project has received funding from the Medical Research Council (MRC). More information about MRC can be found here: https://www.ukri.org/councils/mrc/

#### Participation in future research

Should you wish to be involved in future research your contact details would be held on a password protected computer in room N6, ITTC building, Plymouth science park. Agreeing to be contacted does not oblige you to take part in future research.

If you are interested in finding out more about getting involved in research visit: https://www.nihr.ac.uk/patients-carers-and-the-public/

#### Further information and contact details

Please contact Dr William Heseltine-Carp at or Dr Stephen Mullin at

Alternatively, you can write to:

Dr Stephen Mullin
Room N6
ITTC Building Plymouth Science Park
Derriford
Plymouth
PL6 8BX

*Thank you for considering taking part.*
