## Appendix 2 for "Opinions of the UK general public in using artificial intelligence and “opt-out” models of consent in medical research"

### Focus group discussion of use of routine data - pre read documents

### How do we ensure our research is ethical?

Each project will be based on a defined research question or objective. Careful consideration will have been given to what the minimum amount data required to robustly answer this question. The scientific validity of this question and the need for the use of such data will in every case be considered by a research ethics committee. Only research which seems to be able to deliver direct benefits to patients will be approved. This a panel of doctors, scientists and members of the public who consider whether the proposed research is ethically justifiable.

The project that we are working on at the moment is called '*Using Explainable Artificial Intelligence to Predict Future Stroke using Routine Historical Investigations'.*

In this project we aim to use the results of medical tests that people normally have when admitted to hospital (for example, blood tests, heart scans and brain scans) to predict someone's risk of having a stroke in the future.

Eventually we might be able to use this information to identify people who might be at high risk of stroke and provide medical intervention to prevent them from having a stroke in the future.

We also hope this research will help us identify new risk factors for stroke and therefore ways in which we might be able to lower peoples risk of having a stroke.

To do this we plan on using the data of approximately 10,000 people who have had a stroke in the past and 80,000 people who haven't had a stroke. Because of these large numbers it would be impractical to gain consent from all these peoples individually.

That's why we will be using people's data in this research without their elicit consent. To make sure we're doing this in an ethical way, we plan on anonymising their data in our research, protecting their identity.

This is why we have invited you to our focus group. We want to make sure that we have gone to enough steps to ensure that our research is ethical and that the identity of people involved in our research is protected.

You can find more information on this research project in the invitation pack above.

### Questions for the day

Below you can find 3 questions that we plan on discussing on the day. Each question aims to address a potential issue we might have when performing research in this way.

Below each question includes information on how we plan to address each issue. Have a read of this information and think whether the steps that we have taken are enough to address this issue the question is highlighting.

**1) Is it acceptable to collect, store and analyse data in this manner for medical research within?**

*When research like this is performed data is often accessed by two different groups of researchers. This can be divided into researchers within the routine clinical care team, and researchers outside of the routine clinical care team.*

#### ***Data collection***

*Within routine clinical care team*

*The routine clinical care team refers to people who normally provide clinical care for an individual and would routinely have access to their data. This might include doctors and nurses part of the medical team that have looked after them in hospital. Wherever it is possible we will ensure that your data is handled only by members of the routine clinical care team.*

*Outside the routine clinical care team*

*Sometimes this is not possible, for instance when data must be extracted from different hospitals in order to produce a large enough group of participants to answer a research question. In this case the data will remain within NHS systems, but would be processed by NHS staff who would not ordinarily have access to it. This is known as transfer of identifiable data 'outside of the routine care team'. This might include doctors and nurses who work in the NHS, but have not been part of the clinical team providing clinical care for the individual.*

#### ***Data storage and de-identification***

*When we collect data like this we only use it for research purposes. We also try to make sure that when we collect data it is done in an ethical way, making sure we protect the identity of anyone included in the dataset. There are a number of ways we ensure this, which are described below.*

*Storage and de-identification of data*

*In order to ensure that identifiable data (that is data which contains information which may be used to identify participants) remains confidential, this type of data will only be handled on NHS IT systems by NHS staff. A number of steps are required to produce datasets for this work*

*Subject identification*
*People with stroke specific medical problems of interest (in this case stroke) will be identified from hospital records. This will be done by the 'routine clinical care team' (defined as above).*
*A list of ' matched controls controls' will also be produced. These are people who do not have stroke, but have had the same tests as someone with stroke, for example blood tests or brain imaging.have a medical condition.*
*Comparing the results in those who've had a stroke against those who haven't had a stroke might help us better understand what causes stroke, who is at high risk of stroke and how we can prevent people from having stroke.*

*Linkage*
*People's test results are often held on different databases. For example, some information may be held with your GP and other with a hospital. We therefore need to combine this information from different sources into one database. This process is known as 'data linkage'.*
*To do this we will use your 'NHS number' (a unique number given to everyone in the UK under the care of the NHS). Your NHS number will be used to link information such as blood results, radiology scan pictures and reports (CT and MRI), hospital records and GP records from different databases, to one single database.*

*De Identification*
*During creation of this single database, all information which could be used to identify participants will be removed from the database. This is known as 'de-identification'. This would include information such as your name and home address. The database will be audited to ensure no identifiable information remains. Only once the database is de-identified and anonymised will it be released for researchers outside of the NHS for analysis. This process ensures that your data remains protected and that no information could be traced back to you.to analyse.*

**2) Has sufficient opportunity for participants to 'opt out' of research has been given?**

*Opting out*

*Because we know that not everyone will be comfortable having their data used in this way, we have made it possible to opt out of such research.*

*National opt out*
*Before people are selected to take part in the study, participants who are identified are cross checked against the NHS England 'data opt-out register'. This national database records everyone who has requested their data not be shared outside of the routine care team.*

*Local opt out*
*We also provide a facility for 'local opt out'. This means we will advertise and promote the studies within the region (and nationally if data is being collected outside of the South West Peninsula) through public engagement events. We will offer the opportunity for people to opt out of individual projects or all projects run by our research groups using a portal on our website.*

**3) Have you any questions or further points you would like to add?**

*This might include additional outcomes that we should measure (for example the quality of life after stroke), other areas of research you think are important, how this research could be used in the future to benefit stroke care?*
